# On-scalp magnetoencephalography can detect mesial temporal lobe epileptiform discharges

**DOI:** 10.1101/2023.10.03.23296442

**Authors:** Odile Feys, Maxime Ferez, Pierre Corvilain, Sophie Schuind, Estelle Rikir, Benjamin Legros, Nicolas Gaspard, Niall Holmes, Matthew Brookes, Vincent Wens, Xavier De Tiège

## Abstract

The validation of on-scalp magnetoencephalography based on optically pumped magnetometers (OPM-MEG) for clinical applications requires the assessment of the spatial concordance between reconstructed neural sources and intracranial data recorded simultaneously.

We here report unprecedented data from an epileptic woman suffering from refractory focal epilepsy who underwent a 10-min simultaneous whole-scalp-covering video-OPM-MEG (118 channels) and stereo-electroencephalography (SEEG) recordings.

The most active irritative zone involved the right amygdala despite multiple seizure-onset zones. The multimodal recording allowed to detect and localize 31 simultaneous interictal epileptiform discharges (IEDs) in OPM-MEG signals among the 52 IEDs detected in SEEG signals, and corresponded to the IEDs with the highest amplitude in SEEG. These IEDs were located at the right amygdala using both modalities, with sources distant from 14.1mm.

This simultaneous recording demonstrates the ability of OPM-MEG to detect and localize IEDs from a deep mesiotemporal structure such as the amygdala with a localization value and an IED detection rate in line with previous simultaneous cryogenic MEG and SEEG studies. It paves the way for further on-scalp OPM-MEG recordings in patients with implanted materials and strengthens the potential of this growing technology for future clinical and research applications in patients with brain disorders.

## Introduction

Cryogenic magnetoencephalography (MEG) contributes to the detection and localization of epileptiform discharges in patients suffering from refractory focal epilepsy. In such systems, magnetic field sensors require cryogenic cooling and are housed in a one-size-fits-all, commonly adult-sized, helmet to maintain thermal isolation (2–4 cm) from the scalp. This reduces the signal-to-noise ratio (SNR) and adaptability to the pediatric population ^1^. Cryogenic MEG has demonstrated its ability to detect and localize epileptiform discharges emanating from deep brain structures such as the amygdala or the hippocampus thanks to simultaneous recordings with stereo-electroencephalography (SEEG) ^2^.

Optically pumped magnetometers (OPMs) are novel cryogen-free magnetic field sensors. They have enabled the development of revolutionary on-scalp MEG systems that are movement ^3,4^ and lifespan ^5^ compliant. By bringing sensors closer to the brain, on-scalp OPM-based MEG (OPM-MEG) should also lead to a substantial increase in neuromagnetic field amplitude and SNR. Case studies demonstrated the ability of OPM-MEG to detect and localize ictal ^4^ and interictal ^6,7^ epileptiform discharges from multiple neocortical areas with similar or higher amplitude and SNR compared with cryogenic MEG. None of these studies examined the sensitivity of OPM-MEG for the detection of epileptiform discharges originating in mesiotemporal areas. Two case studies relying on a hippocampal-dependent cognitive task ^8,9^ demonstrated the ability of OPM-MEG to detect and localize hippocampal task-related theta power changes in healthy subjects. In most cases, the studies compared OPM-MEG with subsequent, separate cryogenic MEG or intracranial stereo-electroencephalography (SEEG) recordings and are thus biased by the absence of simultaneous recording.

Here, we describe a simultaneous SEEG and OPM-MEG recording that investigates the ability of OPM-MEG to detect and localize interictal epileptiform discharges (IEDs) from mesial temporal lobe structures.

## Methods

A 20-to-30-year-old woman with drug-resistant focal epilepsy associated with a large right temporo-parietal cortical malformation and multiple homolateral nodular heterotopias underwent a 10-min simultaneous video-OPM-MEG-SEEG recording at the end of her two-week video-SEEG recording. The most active irritative zone on SEEG was located at the right amygdalo-hippocampal complex and multiple seizure-onset zones were identified.

The Institutional Ethics Committee approved the study (Hôpital Erasme, P2020/664). The patient gave written informed consent.

Forty-eight OPMs (118 channels, 20 QZFM-G3 – 28 QZFM-G2, QuSpin Inc; sampling frequency: 1200Hz) were positioned on an adult-sized 3D-printed rigid helmet (Cerca Magnetics Ltd) to conduct whole-scalp-covering, close-to-scalp, OPM-MEG recording ^10^. See ^4^ for noise reduction methods and data acquisition.

Eighteen depth electrodes (186 intracranial contacts, DIXI Medical) were implanted using MEG-compatible screws in the right hemisphere according to the conclusion of the multidisciplinary epilepsy surgery meeting (temporal lobe: 9, parietal lobe: 7, occipital lobe: 2). Sixty-four intracranial contacts (63, grey matter; 1, white matter, intracranial reference) were selected to record SEEG signals (EEG amplifier, Advance Neuro Technology, placed within the magnetic shielded room; sampling rate 2048 Hz, no band-pass filter) simultaneously with OPM-MEG signals (synchronization using a 3-Hz square-wave trigger signal).

Noisy OPM channels were excluded after visual inspection. See ^6^ for OPM-MEG signal preprocessing. SEEG signals were referenced to the white matter contact, downsampled at 1200Hz and band-pass filtered at 0.5-300Hz. IEDs were visually detected (O.F.), first in the OPM-MEG signals (OPM-IEDs, blind to SEEG signals), then in the SEEG signals (SEEG-IEDs). The amplitude of SEEG-IEDs detected and undetected by OPM-MEG was compared using two-sided unpaired *t* test (significance, p<0.05).

OPM-MEG signals were averaged separately at the peak of OPM-IEDs and at the peak of SEEG-IEDs undetected by OPM-MEG. The neural source at the peak of the averaged OPM-IEDs was localized using distributed source modeling (using three axes in QZFM-G3 and two axes in QZFM-G2; see ^6^ for source reconstruction method).

## Results

Eight OPMs (18 out of the 116 channels) were excluded from the analysis due to noisy signals. Thirty-one OPM-IEDs (amplitude: 1.00 ± 0.76pT, SNR: 2.75 ± 2.55) and fifty-two SEEG-IEDs were detected (Figure 1). All OPM-IEDs were simultaneously detected by SEEG. SEEG-IEDs detected by OPM-MEG were of significantly higher amplitude (1918 ± 107 μV/cm) than SEEG-IEDs undetected in the OPM-MEG signals (922 ± 85 μV/cm, p=8.97x10^−10^). Averaging of OPM-MEG signals at peak of SEEG-IEDs undetected in OPM-MEG signals did not evidence any clear IED in OPM-MEG signals. The electrode contacts involved by the highest amplitude SEEG-IEDs and the neural source of OPM-IEDs were 14.1mm apart and located at the right amygdala (Figure 1, no SEEG-IED from other less active irritative zones recorded during the 10-min recording).

**Figure 1.**
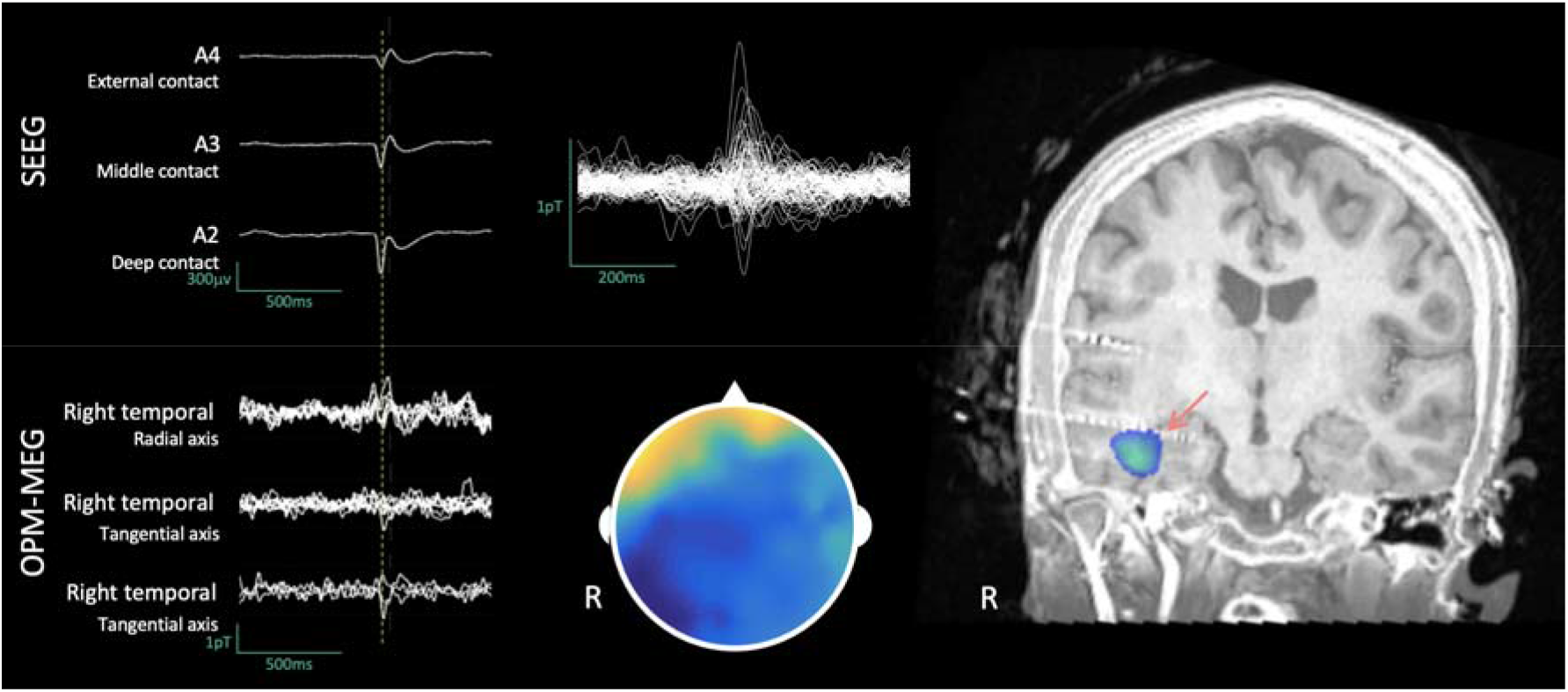
Neurophysiologic and neuroimaging data. **Left**. Samples of simultaneous 1-sec signals of SEEG (band-pass filter: 0.5Hz-300Hz, **Top**) and OPM-MEG (band-pass filter: 3-40Hz, **Bottom**) recordings showing one IED detected by both modalities. **Middle**. Averaged OPM-MEG signals at the peak of OPM-IEDs (**Top**) and corresponding magnetic field topography (arbitrary scale, **Bottom**). **Right**. Coronal slice of co-registered pre-implantation T1-weighted brain magnetic resonance imaging and post-implantation brain computed tomography with the neural source reconstructed at the peak of averaged OPM-IEDs (blue) and with the intracranial SEEG electrode contacts recording IEDs with the highest amplitude (red arrow).

## Discussion

The validation of on-scalp OPM-MEG for clinical applications critically requires the demonstration of a reasonable spatial concordance between reconstructed neural sources and intracranial data recorded simultaneously ^11^. This case study demonstrates, using such simultaneous recording, the ability of OPM-MEG to detect and localize IEDs from a deep mesiotemporal structure such as the amygdala.

The distance between the neural source of averaged OPM-IEDs and the electrode contacts involved by SEEG-IEDs was higher than the ∼5mm spatial resolution of cryogenic MEG but in line with previous simultaneous cryogenic MEG-SEEG studies that recorded mesiotemporal epileptiform discharges (from 8.4 to 29.8mm ^12^). This discrepancy could be related to multiple non-exclusive factors. First, the SEEG spatial brain sampling is intrinsically limited by the number of depth electrodes to be implanted and by the *a priori* hypotheses about the location of epileptogenic zone ^13^. The combination of SEEG with simultaneous whole-scalp-covering MEG recordings allows to overcome this limited spatial sampling of SEEG ^2,12,14^. In this case, no electrode was directly implanted within the reconstructed neural source of OPM-IEDs. It is thus impossible to know which of the reconstructed OPM-MEG source or the depth electrode was the more accurate to locate the neural source of IEDs. Second, multiple coregistration imprecisions (between cerebral structural MRI and post-implantation computed tomography, between cerebral structural MRI and OPM-MEG spaces) can lead to an underestimation of OPM-MEG spatial accuracy. Third, the low SNR of OPM-IEDs may lessen OPM-MEG spatial accuracy.

The SNR of OPM-IEDs was lower than in previous OPM-MEG data that recorded neocortical epileptiform discharges (2.75 vs. 11.1 to 16.7) ^5,6^ but similar to previous simultaneous cryogenic MEG-SEEG studies that recorded neocortical epileptiform discharges (∼2)^14^. This can be related to (i) the deeper location of mesiotemporal structures compared with neocortical areas, (ii) to a higher brain-to-sensor distance due to close-to-scalp rather than on-scalp OPM-MEG recording, or (iii) to the proximity of OPMs with screws/intracranial electrodes, moving cables or the EEG amplifier within the magnetic shielded room. The presence of screws and intracranial electrodes prevented the use of flexible EEG-like caps previously used for on-scalp OPM-MEG recordings^6^ and favored the use of a large 3D-printed helmet to house the OPMs and safely contain the patient’s head with the implanted SEEG electrodes,^10^ leading to a close-to-scalp recording and increased brain-to-sensor distance. Sixty percent of SEEG-IEDs were successfully detected by OPM-MEG, corresponding to those with the highest amplitude. This finding is at the upper end of the 25-60% detection rate of mesiotemporal IEDs in previous simultaneous cryogenic MEG-SEEG studies ^13^.

Our case-study demonstrates the feasibility of wearable OPM-MEG in patients with implanted intracranial devices. It thus opens the door for future on-scalp or close-to-scalp OPM-MEG recordings in adverse recording situations, such as SEEG to overcome its limited spatial brain sampling or implanted neuromodulation devices (e.g., responsive neurostimulation). Specific OPM-MEG denoising methods such as signal space separation ^15^ could increase the IED detection rate of OPM-MEG in patients with implanted intracranial materials.

This case study demonstrates using simultaneous OPM-MEG and SEEG recordings the ability of on-scalp OPM-MEG to detect and locate deep mesiotemporal epileptiform discharges. This finding strengthens the potential of this growing technology for future clinical and research applications in patients with brain disorders.

## Data Availability

Data are available upon reasonable request to the corresponding author and after approval of institutional (Hopital universitaire de Bruxelles & Universite libre de Bruxelles) authorities.

## Funding information

O.F. is supported by the Fonds pour la formation à la recherche dans l’industrie et l’agriculture (FRIA, Fonds de la Recherche Scientifique (FRS-FNRS), Brussels, Belgium). P.C. is supported by the Fonds Erasme (Convention " Alzheimer ", Brussels, Belgium). X.D.T. is Clinical Researcher at the FRS-FNRS. The other authors received no additional funding.

The OPM-MEG project at the Hôpital Universitaire de Bruxelles and Université libre de Bruxelles is financially supported by the Fonds Erasme (Projet de Recherche Clinique et Convention “Les Voies du Savoir 2”) and by the FRS-FNRS (Crédit de Recherche: J.0043.20F, Crédit Équipement: U.N013.21F).

## Competing interests

N.H. and M.B. holds founding equity in Cerca Magnetics Limited, a spin-out company whose aim is to commercialize aspects of OPM-MEG technology. The remaining authors have no conflicts of interest.

## Data availability statement

Data are available upon reasonable request to the corresponding author and after approval of institutional (Hôpital universitaire de Bruxelles & Université libre de Bruxelles) authorities.

## Notes

### Funding Statement

O.F. is supported by the Fonds pour la formation a la recherche dans l industrie et l agriculture (FRIA, Fonds de la Recherche Scientifique (FRS-FNRS), Brussels, Belgium). P.C. is supported by the Fonds Erasme (Convention Alzheimer, Brussels, Belgium). X.D.T. is Clinical Researcher at the FRS-FNRS. The other authors received no additional funding.
The OPM-MEG project at the Hopital Universitaire de Bruxelles and Universite libre de Bruxelles is financially supported by the Fonds Erasme (Projet de Recherche Clinique et Convention Les Voies du Savoir 2) and by the FRS-FNRS (Credit de Recherche: J.0043.20F, Credit Equipement: U.N013.21F).

### Author Declarations

The Institutional Ethics Committee approved the study (Hopital Erasme, P2020/664). The patient gave written informed consent.

